# CULTURAL CONSANGUINITY AS CAUSE OF β-THALASSEMIA PREVALENCE IN POPULATION

**DOI:** 10.1101/2023.06.01.23290856

**Authors:** Muhammad Aslamkhan, Muhammad Imran Qadeer, Muhammad Shoaib Akhtar, Shafiq Ahmad Chudhary, Maida Mariam, Zain Ali, AbdurRauf Khalid, Muhammad Irfan, Yasin Khan

## Abstract

**Background:** Some 200 million people worldwide have haemoglobinopathies of some sort. Pakistan, where 80% consanguinity prevails because of marriages within caste groups that are anthropologically same. The study aims to reveal the impact of consanguinity on thalassemia in various castes in Punjab, Pakistan.

**Subjects and Methods:** 262 β-thalassemic patient’s families were studied. Patients were registered in various Thalassemia Blood Transfusion Hospitals/Centers, in the metropolitan city of Lahore, Punjab, Pakistan. Patients and parents were interviewed using structured questionnaire regarding information about name, age, sex, ethnicity (caste), educational status, consanguinity of parents, number of progenies, health status of children, pregnancy wastage and family history.

**Results:** The 262 (couples) parents of β-thalassemic patients revealed 96% consanguineous marriages with 72% first cousins, 10% distant blood relatives and 14% *Bradari*. Inter-castes marriages were 4% only. These families produced 1646 children, 582 males and 464 females. Of these, 303 boys and 293 girls are healthy, while 279 boys and 171 girls are thalassemic. In 26 castes, the prevalence of thalassemia varied from 21% to 3%. Rajput tribe on top followed by Arain.

**Conclusion:** Thalassemia is widely spread in Pakistani population. Its prevalence varied in caste groups due to endogamy, a major impact on the prevalence of thalassemia.

## Introduction

Many epidemiological studies revealed the association of parental consanguinity with congenital malformations (Aslamkhan et al., 1969, Akhtar et al., 2019, Zar et al., 2020, Afzal et al., 1994, Ullah et al., 2015, Aslamkhan, 2015, Hamamy, 2012). Progeny of consanguineous marriage usually shows many congenital disorders, like neural tube defects, congenital heart defects, epilepsy, and mental retardation (Peyvandi et al., 2002, Madhavan and Narayan, 1991, Hamamy et al., 2007, Fernell, 1998, Mahadevan and Bhat, 2005, Nauman, 2016). According to the World Health Organization about 7% of the World population is carrying haemoglobinopathies that are heritable. It is also estimated that about 5 million infants are born with severe form of congenital diseases annually. Consanguineous marriages are extremely common in Pakistan, South Asia and Muslim world (Tadmouri et al., 2009, El-Kheshen and Saadat, 2013, Akhtar et al., 2019, Zar et al., 2020).

Thalassemia is an autosomal recessive disorder characterized by mutation in regulatory gene that results in under production of globin proteins. Haemoglobinopathies imply structural abnormalities in the globin proteins themselves (Cousens et al., 2010). The prevalence of thalassemia is more common in population living in habitats having humid climates and of malaria endemic area (Salih et al., 2010). This association between thalassemia and humid climates is result of natural selection to protect masses from parasitic infections which are common in humid climate regions (Hedrick, 2011, Qiu et al., 2013). However, its consequences are observed in all races (Shah et al., 2017). The people of Mediterranean origin, Asians and Arab-Americans are mainly suffering from thalassemia (Mazharul Islam, 2017). It is noticed that about 16% people from Cyprus, 3-14% from Thailand, 3-8% population from India, Pakistan, Bangladesh, and China are victims of thalassemia (Kalokairinou, 2007, Riewpaiboon et al., 2010, Asif and Hassan, 2014, Ahmed et al., 2022, Colah and Gorakshakar, 2014, Colah et al., 2017, Hossain et al., 2017, Zhang et al., 2018, Yao et al., 2013). However, the prevalence of thalassemia in Pakistan is not known.

Inherited hemoglobin disorders provoke many clinical sign and symptoms including severe hemolytic anemia, jaundice, recurrent infections, and traumatic crisis leading towards high morbidity, mortality, and fetal wastage of progenies (Trehan et al., 2015). The sufferers involve the newborn babies, growing children, juvenile girls, pregnant women, and a large portion of illiterate population (BORGNA□PIGNATTI et al., 2005). The alliance of consanguinity with thalassemia morbidity and mortality has been advocated (Aslamkhan, 2015, Aslamkhan et al., 1969). Epidemiological studies have a power to look for underlying mechanisms and lead to potential solutions (Akthar, 2019). This study is also an effort to document epidemiology and etiology of thalassemia for developing a control and preventive strategy in isonymic endogamous ethnic tribes living in the Punjab, Pakistan.

## Subjects and Methods

All the children and adolescents braving β-thalassemia, between the ages of 6 months and 20 years, and above, who were registered and receiving blood transfusion from various blood transfusion centers including Children Hospital, Ganga Ram Hospital, Mayo Hospital, Fatimid Foundation, Hilal-e-Ahmer and Sundas Foundation, in the Metropolitan city of Lahore, Punjab, Pakistan, were recruited in this study. The current study was conducted during the period from October 2014, till March 2015. The study protocol was descriptive, irrespective of their ethnicity, caste, or any other social factor. Only the index cases in each identified β-thalassemic potential family were included in this study. The required data including isonym ethnic tribe (caste), age, sex, family detail, genetic history, parental consanguinity, total number of children in the family, number of β-thalassemia afflicted males and females, pregnancy wastage, number of children died, type of β-thalassemia, educational and financial status were collected by the authors through interview (patients and/or their parents). After obtaining their written consent on a structured Proforma. The protocol was approved by the IEC (Institutional Ethical Committee). Parental consanguinity was classified into four groups as: (i) first cousins, (ii) near/distant blood relatives, (iii) Baradari (brotherhood), same caste and isonym ethnicity (intra-caste) but blood relationship not known), and (iv) different caste (inter-caste) marriages.

## Results

### High prevalence of consanguinity among β-thalassemic families

A total of 262 families (parents of β-thalassemic patients) were studied. Consanguinity of parents were observed and reported in following four groups; (i) 72% were first cousins, (ii) 10% were near/distant blood relatives, (iii): 14% were intra-caste, and (iv): 4% were inter-caste marriages (Figure 1).

**Figure 1:**
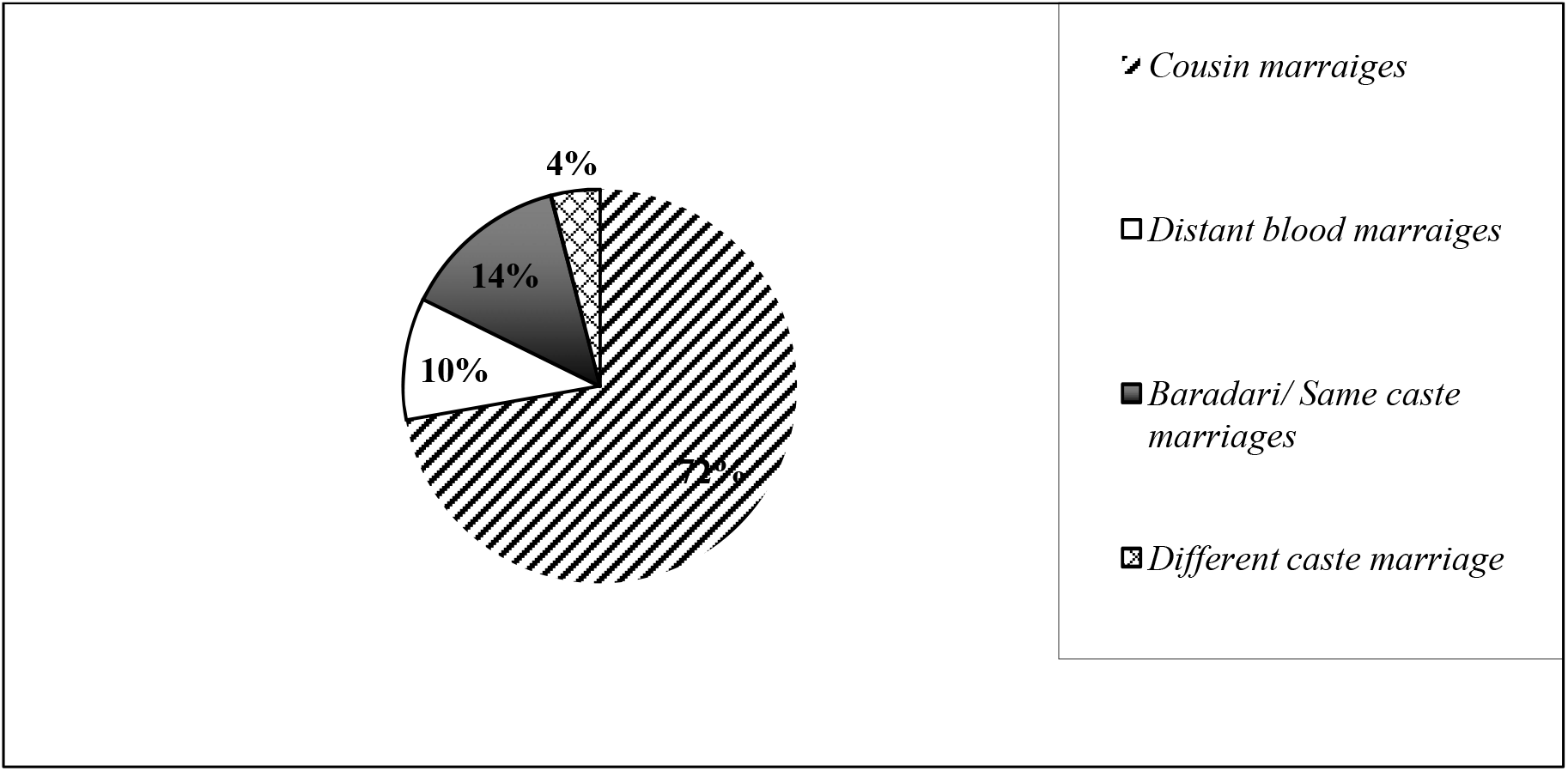
Consanguineous pattern in 262 β-thalassemic studied families

These 262 families produced a total of 1046 children, 582 (55.64%) males and 464 (44.36%) females, having 3.99 progeny per family. Of these 596 (56.12%) children, 303 (52.06%) boys and 293 (63.15%) girls, were healthy while 450 (43.02%), 279 (47.94%) boys and 171 (36.85%) girls) were β-thalassemic. Of these 450 β-thalassemic, 90 (8.60%) were deceased due to thalassemia, 53 males (58.88%) and 37 females (41.11%).

### β-thalassemia distribution by caste

These 262 studied families of β-thalassemic carriers consisted of 26 isonym groups (castes), in which the prevalence of thalassemia varied as follows: Rajput 21.0%, Arain 17.0%, Sheikh 10.0%, Mughal 8.0%, Jatt 6.1%, Pathan 5.3%, Khokhar 5.0%, Malik 3.4%, Kamboh 3.0%, Gujjar 2.6%, and Miscellaneous 19%. Miscellaneous group consists of 16 castes having less than 2% prevalence of β-thalassemia. These tribes include Awan, Ansari, Kakazai, Mahar, Joya, Mukandy, Bangash, Bhatti, Ghumman, Butt, Chohan, Ghaffari, Rehmani, Royal, Sayed and Christian (Figure 2). We observed significant positive correlation between caste and β-thalassemic children in all castes except in Pathan, Kamboh and Malik. (Table 1).

**Figure 2:**
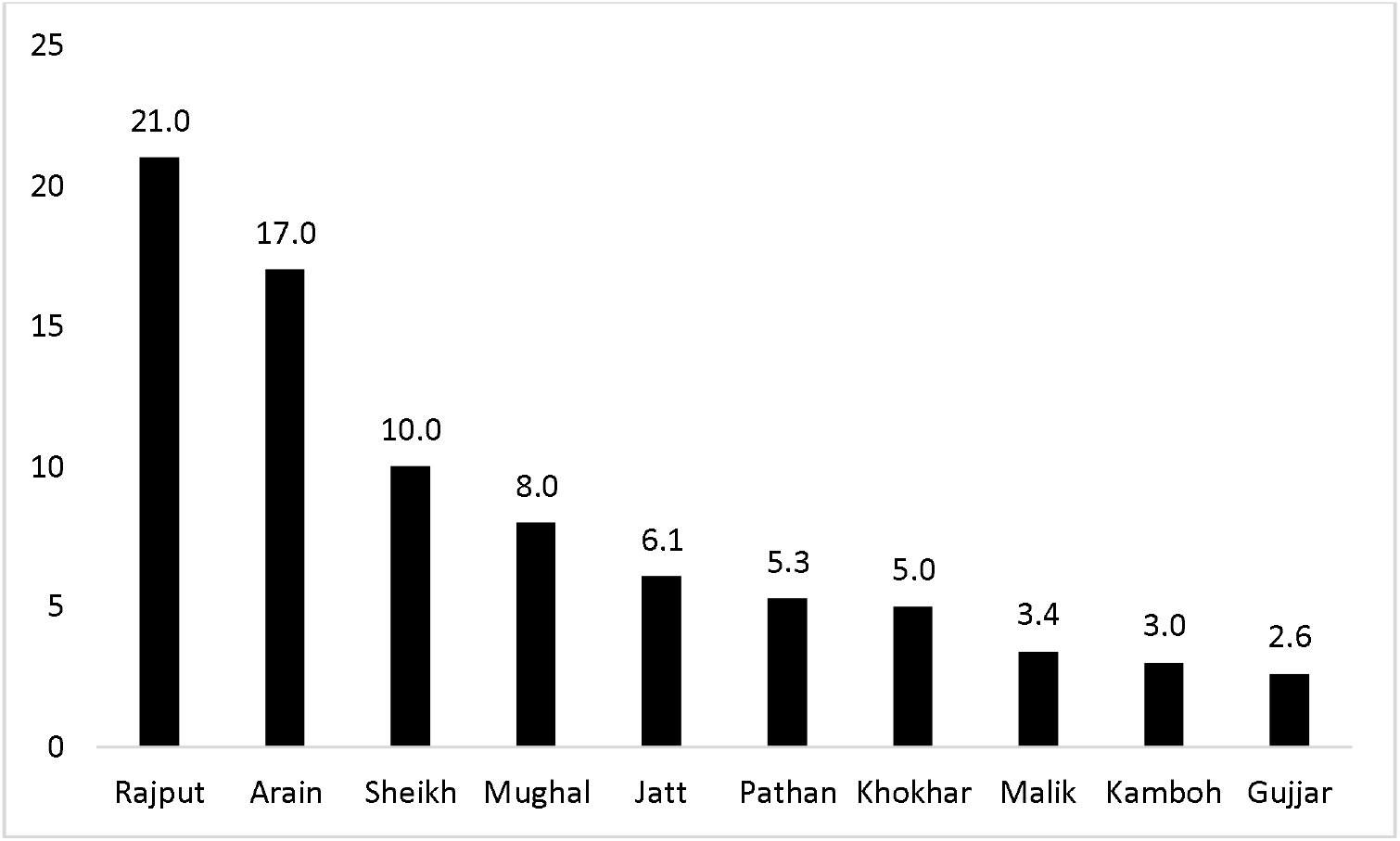
Prevalence of β-thalassemia in various castes in studied population. Castes are shown in in decreasing order by prevalence. 16 castes with prevalence less than 2% are grouped together as miscellaneous and are not shown in the figure.

**Table 1:**
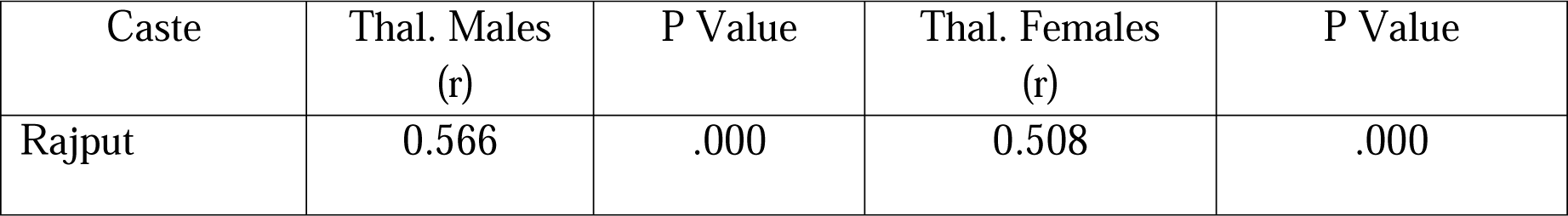
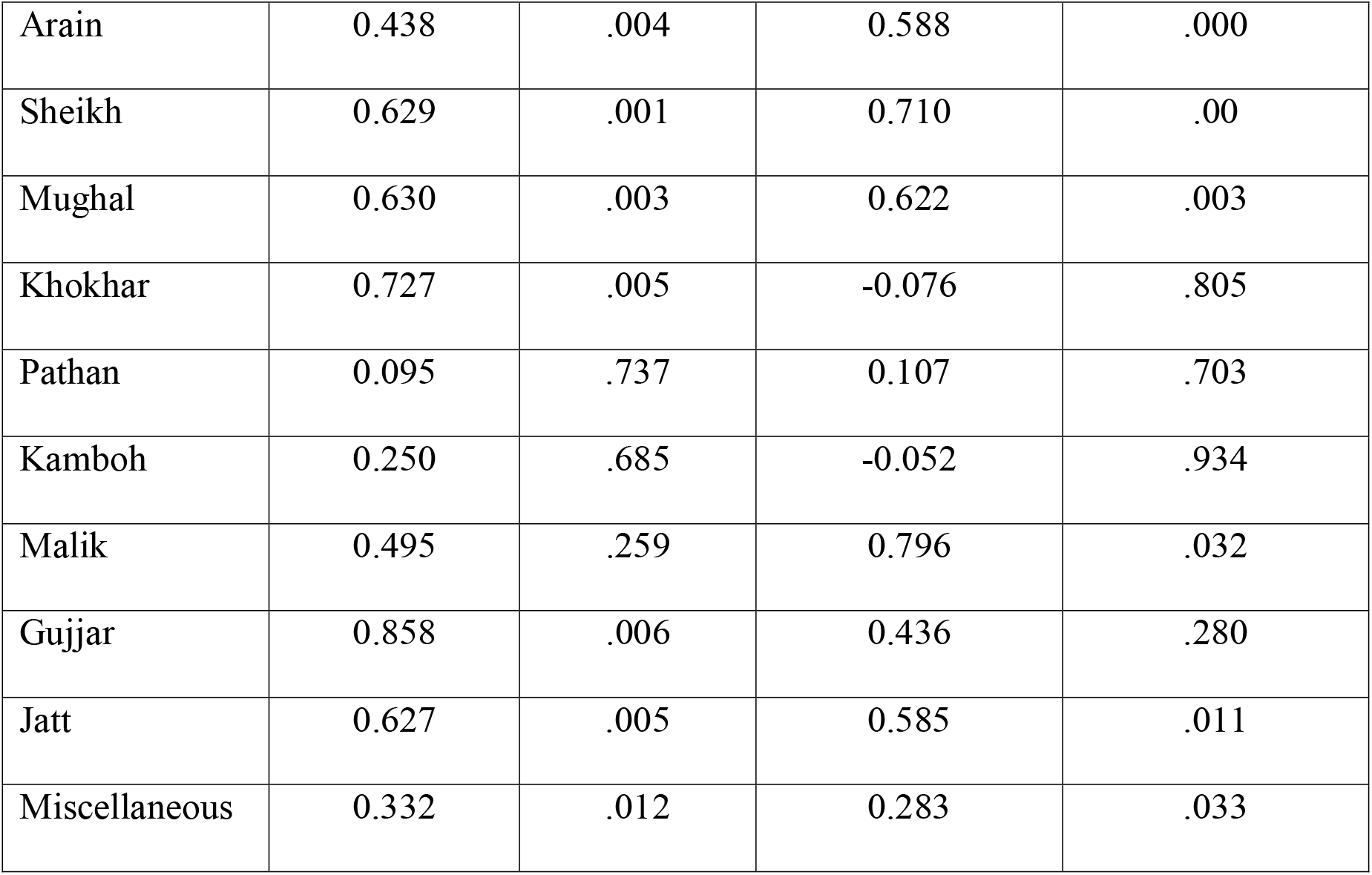
Correlation between the caste and β-thalassemia children in studied families.

These 450 patients, ages between 5 to 20 years, having β-thalassemia minor, intermediate and major were included in this cross-sectional study. The male/female ratio in these children of β-thalassemia carrier was 3:2. The etiology of β-thalassemia was identified in 450 (43.02%) of the children, other 596 (56.12%), apparently normal, were considered to be heterozygous or normal.

### Consanguinity as cause of β-thalassemia

The most common cause of β-thalassemia in these patients was considered to be consanguineous marriages of their parents. 96% parents were married to their cousins, distant blood relatives, or intra-caste (72%, 10% and 14%) as shown in Figure 1. In all marriages, first cousin of maternal lineage is preferred and the caste being the major factor in this regard.

Table 1 tabulates correlation between β-thalassemia and consanguinity. Among first group(I: first cousin marriages), five out of 10 castes showed significant correlation. No significant correlation was observed in group two (II: Near/distant blood relative marriages). But Rajput caste showed significant correlation in first cousin marriages. In third group (III: Bradari/Same caste but unrelated marriages), two castes out of four, arain and sheikh, showed significant correlation. Rajput which showed significant correlation among first cousin marriage group did not show correlation in this group again. Primary reason was lower number of Rajput subjects in group II and III. Forth group which was different caste marriage did not show any correlation because this type of marriages was only seen in miscellaneous groups. As castes in this group had very low number of β-thalassemia patients so it explains lack of correlation observation and role of inter-caste marriages in prevention.

## Discussion

In this study, the 96 % consanguinity (first cousin, distant blood relatives and same caste marriages) among parents of β-thalassemia patients is observed (Figure 1). Prevalence of β-thalassemia in castes with higher consanguinity was also observed higher and a significant correlation was also observed (Figure 2 and Table 1). A significant positive correlation was observed between consanguinity and β-thalassemia patients (Table 2). Caste system was the major factor influencing the cultural consanguinity and inbreeding. Our study indicates that consanguinity is an important potential risk factor for thalassemia (Cunningham et al., 2004, Khan, 1983). This finding indicated that consanguinity is a major risk factor for high rates of prenatal mortality and congenital malformation as opined by Aslamkhan, and others (Khan, 1983, Cunningham et al., 2004). Among Pakistani population the risk of thalassemia due to cultural consanguinity is 2.5 times higher than the children of parents who are not near or distant blood related. The study of Ansari reported 43% thalassemic cases in consanguineous marriages and he opined that relationship of consanguinity and thalassemia appears reasonable (Ansari et al., 2011). Previous studies showed consanguinity prevalence of 80%-100% in Pakistani population (Aslamkhan et al., 1969, Akhtar et al., 2019, Zar et al., 2020).

**Table 2:** Correlation between β-thalassemic children and consanguinity.

| **Table2: Correlation between β-thalassemic children and consanguinity** |  |  |  |  |
| --- | --- | --- | --- | --- |
| **I: First cousin marriages** |  |  |  |  |
| **Caste** | **Thal. Males (r)** | **p value** | **Thal. Females (r)** | **p value** |
| Rajput | 0.58 | 0.000 | 0.562 | 0.000 |
| Arian | 0.378 | 0.043 | 0.563 | 0.001 |
| Sheikh | 0.61 | 0.021 | 0.91 | 0.000 |
| Mughal | 0.551 | 0.051 | 0.695 | 0.008 |
| Khokhar | 0.655 | 0.078 | -0.276 | 0.509 |
| Pathan | -0.038 | 0.922 | -0.105 | 0.788 |
| Kamboh | 0.25 | 0.685 | -0.52 | 0.934 |
| Malik | 0.577 | 0.423 | 0.866 | 0.134 |
| Jatt | 0.647 | 0.009 | 0.605 | 0.017 |
| Gujjar | 0.858 | 0.006 | 0.436 | 0.280 |
| Miscellaneous | 0.292 | 0.089 | 0.179 | 0.303 |
| **II: Near/distant blood relative marriages** |  |  |  |  |
| Rajput | 0.408 | 0.495 | -0.134 | 0.83 |
| Moghul | 0.775 | 0.225 | 0.816 | 0.184 |
| Miscellaneous | 0.486 | 0.222 | 0.905 | 0.002 |
| **III: Bradari/Same caste but unrelated marriages** |  |  |  |  |
| Rajput | 0.894 | 0.106 | -0.258 | 0.742 |
| Arian | 0.688 | 0.028 | 0.628 | 0.052 |
| Sheikh | 0.718 | 0.045 | 0.494 | 0.214 |
| Pathan | 0.5 | 0.667 | -0.5 | 0.667 |
| Miscellaneous | ----- | ---- | 0.686 | 0.041 |
| **IV: Different caste marriages** |  |  |  |  |
| Miscellaneous | 0.983 | 0.003 | -0.167 | 0.789 |

In the present study we found that consanguineous marriages were more common among first cousins in maternal lineage (72%) where significant positive correlation is observed between castes and thalassemic children and this disorder was found to be more frequent in families having low educational and social status. Consanguinity undoubtedly increases the birth prevalence of autosomal recessive disorders in multiple consanguineous marriages in thalassemic families resulted in the affliction of males and females. Carrier heterozygous person without manifestation of disease, on marriage show a 25% recurrence risk of homozygosity. The genetic heterogeneity (carriers) having 50% chance of producing more carrier heterozygotes through inbreeding. The chance of inheriting similar two recessive alleles from parents is only 25% thus, this process will keep on increasing the heterozygosity and decreasing the homozygosity, which indicates that consanguineous marriages, either in maternal or paternal lineage, are potentially associated with recessive gene expression in consanguineous progeny (Benson, 2005).

It is important for genetic counsellors to take detailed family history to ascertain the presence of hereditary disorders. This information will help them in genetic counseling and refer them to a healthcare center. It is also possible that in isolated consanguineous marriage couple there may be more than one or two recessive genes or other mutants, which will greatly complicate the counseling (Bittles, 2001). However, as several studies in different parts of the world indicate population screening of extended families and young population of high-risk groups yields good results in the prevention of marriage of two carriers and/or allow opting for antenatal diagnosis. The treatment and management of such genetic disorders greatly enhance the economic burden on the family resources (Weatherall and Clegg, 2001). National government or international health organizations may initiate a pre-marital genetic screening programs in this scenario.

A major shift in the balance between the social and economic benefits associated with intra-caste marriages and adverse health outcome can be predicted especially in affected families. Majority of the couples (85-90%) still come retrospectively after having one or more affected children. A diagnostic, counseling and management skills are urgently needed along with the community awareness programs. Reasons of preferring marriage with blood relatives is deep rooted in the culture of South Asia. In India, the population is divided into caste groups according to the *“Verna”* (means color) philosophy of *Rig Veda*. We are told that Aryan from Central Asia invaded northwest India, circa 1800 BCE, annihilated indigenous tribes and enslaved them. As a result of indigenous tribe admixture, however, production of dark colored progeny with black hair and broad nose alarmed the fair colored, blond hair Aryans to invoke the *Varna* policy, branded the indigenous dark-colored people as untouchable and banned marriage contract with them.

Aryan had no caste but consisted of three groups of people: Religious Elders (*Brahmana*), Warriors (*Khashtria*) and Artisans (*Vaisha*). With the passage of time the ruling groups produced hundreds of power groups, that became known through the name of their founding father. His descendants and followers started using that name as sur name. Each isonym group called a caste and mate selection confined to that caste. Indigenous people given the lowest caste group as “s*chuder”*. They are nicknamed as “defeated”, “coward”, “low”, “black”, “slave” and branded as untouchable. Hindus after conversion to Islam kept the same caste sur name while the indigenous tribe converter adopted the respectful title “Sheikh”. During British Raj, indigenous castes on conversion to Christianity were called as “Anglo-Indian”. Thus, many new cultural isonym groups, e.g., Ghaffari, Rehmani, Royal, Christian, etc. were came into being. Being in minority, these social groups also married with cousins/near blood relation, which became a cultural practice (Aslamkhan, 1996).

Other, so called, advantages of cultural consanguinity advocated by conservative people include: a) cousin marriages bound family through renewed relationship among members and strengthen the family socially and politically; b) cousin marriages enhance family hierarchy in the community; c) it provides suitable partner for both boy and girl who are chosen by the elders; d) consanguinity preserves the family lineage and its prestige; e) it preserves the tradition of the family and its culture; f) cousin marriages keep the land property within the family; g) it keeps the finance of the family intact; h) cousin marriage keeps the status of the girl intact or may enhance it further; i) consanguinity preserves communication between the family intact; j) family emotional issue are not provoked (El-Kheshen and Saadat, 2013, Bittles, 2001, Ahmed et al., 2002).

## Conclusion

The study has revealed the impact of cultural consanguinity which caused 96% β-thalassemia in children of first cousin, distant blood relative and intra-caste marriages versus only 4% in different caste marriages. The incidence of thalassemia varies in different castes, but the actual prevalence of β-thalassemia gene(s) is not known, which requires epidemiological studies to ascertain the prevalence of heterozygote (carrier) of β-thalassemia gene. Parents of β-thalassemia children coming for health care of their children should be encouraged and convinced to test all children in their extended families. Genetic screening of heterozygotes should be mandatory not only for suffering families but also for all children. Detection using molecular techniques of inherited recessive genetic disorders in the population will help in controlling the disease.

## Data Availability

All data produced in the present study are available upon reasonable request to the authors

## Acknowledgement

We thankfully acknowledge the permission given by the Medical Superintendent of various institution to give us excess to the β-thalassemia patients.

## Author Contributions

### Conceptualization

Muhammad Aslamkhan and Muhammad Shoaib Akhtar.

### Analysis

Muhammad Imran Qadeer, Maida Mariam.

### Funding acquisition

Yasin Khan.

### Investigation

Maida Mariam, Shafiq Ahmad Chudhary, Zain Ali, Abdurrauf Khalid and Muhammad Irfan

### Project administration

Muhammad Aslamkhan, Muhammad Imran Qadeer.

### Resources

Muhammad Aslamkhan, Muhammad Imran Qadeer, Maida Mariam, Shafiq Ahmad Chudhary.

### Supervision

Muhammad Aslamkhan, Muhammad Imran Qadeer.

### Visualization

Muhammad Aslamkhan, Maida Mariam.

### Writing – original draft

Maida Mariam and Muhammad Shoaib Akhtar

